# Health system response to climate-related shocks in a lower-middle-income country setting: a comparative study of floods and droughts in Pakistan

**DOI:** 10.1101/2025.11.26.25341133

**Authors:** Marianne Moussallem, Shehla Zaidi, Kate Gooding, Rabia Najmi, Nousheen Pradhan, Ibrahim R. Bou-Orm, Sophie Witter

**Affiliations:** Higher Institute of Public Health, Saint-Joseph University of Beirut, Lebanon; ReBUILD research consortium; University College London, United Kingdom; Aga Khan University; Oxford Policy Management Limited, United Kingdom; Liverpool School of Tropical Medicine, United Kingdom; Queen Margaret University Edinburgh, United Kingdom

## Abstract

Climate-related shocks increasingly threaten population health in Pakistan, where both rapid-onset floods and slow-onset droughts disrupt essential services and heighten vulnerabilities. Despite an expanding disaster management architecture, the extent to which health systems can anticipate, withstand, and adapt to these shocks remains unclear. This study examined how planning, resourcing, coordination, and service delivery function during floods and droughts, and how communities cope with these pressures across high-risk districts.

An exploratory qualitative design was used, drawing on interviews with national, provincial, district, and facility stakeholders; focus group discussions with community members and lady health workers; and a document review. The analysis was guided by the conceptual framework for shock-responsive health systems developed under the Maintains program.

The findings show that while Pakistan has established national and provincial disaster management structures, responses remain largely reactive and uneven across shock types. Floods receive comparatively stronger attention, including annual contingency planning, temporary medical camps, and emergency workforce deployment. However, service disruption remains widespread, particularly for routine care, maternal services, and immunization, and reporting systems become fragmented. Droughts expose deeper systemic weaknesses, including geographic remoteness, poor infrastructure, limited outreach, chronic undernutrition, and minimal anticipatory planning. Across both shocks, coordination between disaster and health agencies is inconsistent, financing is triggered late, and essential service packages lack standardization. Communities demonstrate considerable resilience through local solidarity, informal support networks, and small-scale innovations, yet these coping mechanisms are fragile and insufficient substitutes for reliable state support.

The study highlights the need to shift from reactive responses toward integrated resilience planning across all phases of shocks, with stronger operational planning, clearer role alignment, standardized service packages, and proactive community engagement to protect essential health services during climate-related events.

## Introduction

Climate change presents diverse and escalating threats to human health, altering environmental and social conditions and demanding a robust, coordinated health systems response [1]. Climate change affects health through both direct and indirect pathways. Direct impacts include excess heat-related mortality and morbidity from heat exhaustion and heat stroke, as well as casualties such as drowning during floods or injuries from cyclones and landslides. Indirect impacts operate through a range of environmental and social mechanisms: climate-induced changes in temperature, precipitation, deforestation, and pollution alter the patterns and seasonality of vector-borne, foodborne, and waterborne diseases [1–9]; societal stressors such as mass displacement, disruption of livelihoods, and food insecurity exacerbate existing vulnerabilities, contributing to undernutrition, infectious disease, and mental illness [1,10–12]. Climate-related shocks such as floods, droughts, and landslides amplify these indirect pathways by damaging infrastructure and supply chains, displacing health workers, and straining health systems at a time of heightened demand for services. The burden of both direct and indirect health impacts falls disproportionately on low- and middle-income countries and vulnerable groups, including people living in poverty, children, older adults, and individuals with pre-existing medical conditions [1].

In this study, we focus on health system resilience to two climate-related shocks - floods and droughts - both of which pose significant risks to population health and health service delivery. The World Health Organization (WHO) defines a climate-resilient health system as one that can anticipate, respond to, cope with, recover from, and adapt to climate-related shocks and stresses while continuing to provide essential health services and safeguard population health [1]. Building on this, recent research highlights that health systems in fragile and shock-prone contexts must also be able to absorb the immediate impact of crises, adapt their functions to changing conditions, and, where necessary, transform in order to sustain long-term improvements and progress towards universal health coverage [13,14].

Pakistan is highly prone to climate-related shocks and experiencing an increasing frequency and intensity of events such as floods, earthquakes, heavy rains, landslides, avalanches, and droughts [15–18]. In 2010, the country experienced unprecedented catastrophic floods that displaced millions, caused significant fatalities, and severely damaged infrastructure, including health facilities [11,16,17]. Since then, annual flash floods and urban flooding - driven by glacier melt and increasingly intense monsoon rains - have become common across many provinces. At the same time, prolonged droughts continue to severely affect regions such as Sindh and Baluchistan. These shocks have had major health consequences, including outbreaks of waterborne and vector-borne diseases, widespread service disruption, and serious gaps in maternal health care, especially during major shocks such as the 2022 floods [13–16,19].

In response, Pakistan has established institutional mechanisms including the Ministry of Climate Change and the National Disaster Management Authority (NDMA) with its provincial counterparts, backed by domestic and international investments. However, evidence suggests that responses remain reactive and fragmented, with limited integration into health system planning and weak coordination across levels [15,16, 20, 21]. Broader vulnerabilities such as poverty, weak governance, and limited livelihoods exacerbate these challenges [11].

A growing body of research has examined health system capacity and governance for climate resilience in Pakistan. Yet, this work has often concentrated at the national or provincial policy level. Less is known about how resilience is operationalised at the frontline of service delivery and within communities most directly affected by climate shocks. Detailed evidence from high-risk districts on how health systems prepare, respond, and sustain essential health services during both rapid-onset (floods) and slow-onset (droughts) shocks remains relatively limited.

This study addresses that gap by examining health system resilience in Pakistan through the comparative lens of floods and droughts. It unpacks how planning, resourcing, and coordination shape the continuity of essential health services; identifies critical capacity gaps at subnational and community levels; and explores the coping mechanisms that define community resilience in affected districts. By contrasting the differing demands of flood and drought response, the study contributes new insights to the emerging field of health system resilience to climate shocks.

## Methods

### Study design

An exploratory qualitative study design was used guided by the conceptual framework for shock-responsive health systems developed under MAINTAINS - a multi-country research programme focused on sustaining essential services during shocks and disasters. This framework highlights the importance of health system systems hardware (for example, human resources, supplies, financial resources, and surveillance and information systems) and software (for example, coordination and informal institutions or cultural norms), while also emphasizing the influence of the wider political and governance context, cross-sectoral linkages beyond health, and the role of communities in shaping system responses [22].

The study combined (1) semi-structured key informant interviews (KIIs) with stakeholders from the health and nutrition sectors at national, provincial, district and health facilities’ levels; (2) focus group discussions (FGDs) with community members and community-based Lady Health Workers (LHWs); and (3) review of relevant literature, national programmatic documents and existing government Management Information Systems (MIS) data to situate the findings within the broader context.

### Selection of districts

Districts for this study were selected through a purposive sampling approach-based vulnerability to floods or droughts and inclusion of different health system contexts (Table 1). The aim was to capture a sample of districts affected by floods and droughts, allowing for analysis across varied contexts in Pakistan. Specific selection criteria included:

- Disaster risk vulnerability and geographic diversity: To explore variations in shock impact and response across Pakistan’s diverse settings, very high- and high-risk districts for floods and droughts were selected from different provinces, enabling cross-regional comparisons. Disaster risk vulnerability was based on the most recent classification by the NDMA. Identified districts were then shortlisted by areas that could be accessed within given security clearances, hence excluding high risk districts of Baluchistan and western Sindh [23].
- Recent disaster experience: Priority was given to districts that had experienced major floods or droughts in the two years preceding data collection, ensuring relevance and recency of shock exposure.
- Health and socio-economic indicators: Further shortlisting of districts was based on having weaker health and socio-economic performance, specifically lower immunization coverage, lower skilled birth attendance, and higher rates of stunting compared to other districts in the respective provinces [24–26]. Focusing on low-performing districts helped standardize the sample to areas with existing health system access challenges that are likely to be further exacerbated during climate shocks.

**Table 1.** Table showing district selection.

| Districts | Disaster type | NDMA Hazard Category [28, 29] | Health & Nutrition Indicators |  |  | Province |
| --- | --- | --- | --- | --- | --- | --- |
|  |  |  | Fully immunized children: 12-23 months [30] | Skilled birth attendance [31] | Stunting (moderate and severe) [32] |  |
| Shangla | Flash floods | Very high | 75% | 29% | 78.7% | Khyber Pakhtunkhwa |
| Dera Ghazi Khan | Flash floods | Very high | 92% | 20% | 46% | Punjab |
| Thatta/ Sujawal | Riverine floods | Very high | 30-44% | 49% | 60% | Sindh |
| Larkana | Riverine floods | Very high | 57% | 58% | 52% | Sindh |
| Tharparkar | Drought | High | 79% | 16% | 63% | Sindh |
| Rahim Yar Khan | Drought | High | 84% | 42% | 46% | Punjab |

Within each district, high-risk tehsils and union councils were identified in close collaboration with the Deputy Commissioner (DC) Office and the District Health Office (DHO). A *tehsil* serves as the fundamental unit for general administration, including responsibilities related to treasury, land revenue, land records, and other administrative functions. It maintains the closest and most extensive contact with the rural population. The *union council* plays a key role in local government administration and represents the villages within its jurisdiction [27].

The final sample of districts included Shangla (Khyber Pakhtunkhwa), Dera Ghazi Khan (Punjab), Sujawal and Larkana (Sindh) which are flood-prone, and Tharparkar (Sindh) and Rahim Yar Khan (Punjab) as drought-prone districts.

### Data sources and study participants

A targeted desk review was conducted to contextualize the study and support triangulation of interview findings through review of programmatic documents from government agencies at national, provincial, and district levels and from United Nations (UN) bodies. These included planning documents, guidance and toolkits. Documents were retrieved through targeted website searches and solicitation through interviews. Also, the researchers used secondary data from existing government MIS. The primary source was the District Health Information System (DHIS), which compiles routine data reported from public sector primary and secondary health facilities at the district level. Reporting facilities submit standardized monthly forms covering a wide range of indicators, including outpatient consultations, maternal and child health services, laboratory testing, inpatient admissions, and selected causes of morbidity and mortality. These reports are aggregated quarterly by DHOs. For the flood analysis, Sujawal district was analyzed using the flood period of July–September 2015 and the flood-free period of July–September 2019. For drought, data from Tharparkar district was used, comparing drought period figures in 2018 with a drought-free reference period in 2019. Supplementary material 1 provides information on the documents guiding disaster response reviewed as well as identified areas for consolidation.

#### S1. Documents guiding disaster response and areas for consolidation

In addition to informing the study’s analytical framework, the desk review findings guided the development of data collection tools and provided a reference point for interpreting the primary data.

KII and FGD participants were selected purposively, ensuring representation across national, provincial, district, health facilities and community levels. Initial interview informants particularly national, provincial and district stakeholders were identified through document review and expert consultation, with snowball sampling subsequently employed to expand the sample and capture a broader range of perspectives based on referrals from the first wave of interviews. At facility level, healthcare and nutrition providers were recruited from both primary and referral care facilities across public and private sectors in each district. Two referral facilities that had served as key points during natural disasters were purposively selected per district, with efforts made to include at least one private sector facility. FGDs were held with LHWs and community-based members to understand community-level experiences, coping strategies, and challenges in service delivery during floods and droughts. A list of LHWs and the villages they covered was obtained for each of the selected union councils. Typically, each union council comprises 10–30 villages, with one LHW assigned per village; however, less than half of the villages are usually covered by LHWs. From these listings, 4–6 LHWs per union council were randomly sampled to participate in the FGDs. As for other community members, they were recruited from purposively selected villages in high-risk union councils. Within these villages, participants for FGDs were identified to ensure diversity of perspectives, including men and women, different age groups, and local stakeholders such as elected representatives, elders, and teachers.

KIIs and FGD participants categories are presented in Table 2. In total, the study involved KII with 14 national-level stakeholders, 81 provincial stakeholders, 63 district-level stakeholders, and 63 health and nutrition service providers. In addition, approximately 14 FGDs were conducted with LHWs, and 31 FGDs with male and 34 with female community members.

**Table 2.**
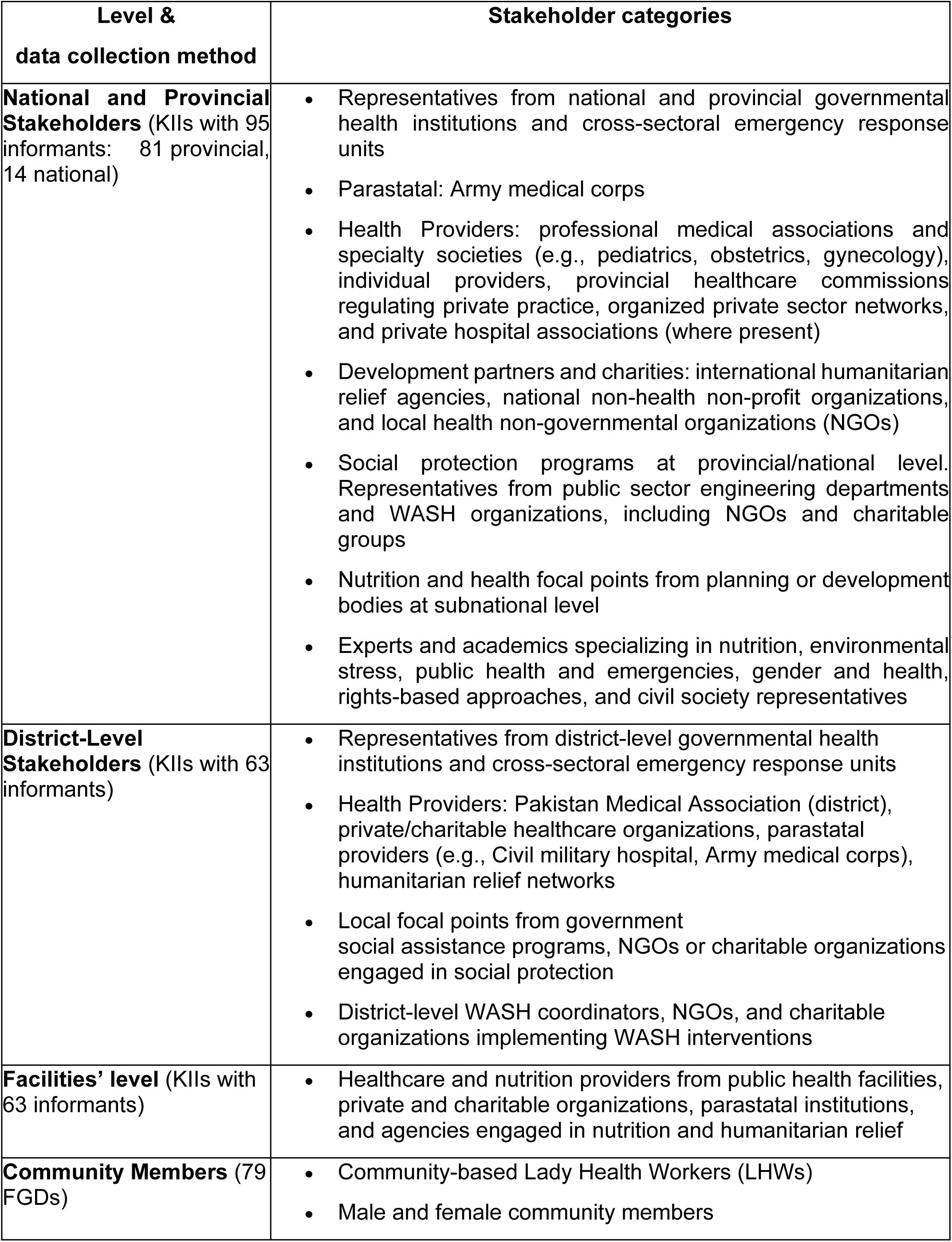
Stakeholders’ categories per level and data collection method.

### Data collection tools and study framework

This study was guided by the MAINTAINS program’s framework [33] (See S2) emphasizing the health system’s ability to maintain routine service delivery while addressing the changing needs during and after shocks - such as epidemics, natural disasters, or conflict. It integrates three interdependent systems: the formal health system (including hardware like infrastructure and workforce, and software such as leadership, trust, and values), the community health system (encompassing community health workers (CHWs), informal providers, and community structures), and other connected systems (such as social protection, WASH, and education). It accounts for both planned and adaptive resilience, acknowledging that systems may absorb, adapt, or transform in response to shocks. Importantly, it underscores the role of governance, institutional trust, community engagement, and intersectoral collaboration, recognizing health systems as complex, adaptive social systems embedded in broader societal and political contexts. Although the model is structured around four phases - preparedness, response, recovery, and reform - which may overlap in practice, this study has mainly focused on preparedness and response phases.

#### S2. MAINTAINS program’s framework

Data collection tools for KII and FGDs were semi-structured and developed based on the MAINTAINS conceptual framework and refined after initial document review.

National and provincial levels’ KII guides focused on strategic insights, and covered the different building blocks, policy formulation, disaster preparedness planning, coordination mechanisms, resource allocation, intersectoral linkages, and reflections on past experiences. Guides for district stakeholders and service providers focused on operational insights, and explored issues such as district preparedness, health workforce mobilization, supply chain readiness, budgeting and resourcing practices, emergency response implementation, and coordination challenges on the ground.

These topic guides also included issues related to the provision of facility-based and outreach nutrition-specific services, information systems used for reporting on disease outbreaks and service utilization, availability of emergency referral systems and coordination with other facilities. For health facility staff specifically, the topic guides were further tailored to assess facility-level preparedness and response capacity, examining infrastructure status, human resource adequacy, including training needs, availability of essential medicines and supplies, referral mechanisms, and continuity of essential health services during and after disaster events.

FGD guides were developed to explore community-level experiences. For LHWs, the tools focused on challenges in service delivery during climate-related shocks, changes in health and nutrition needs, identification of at-risk groups, and suggestions for improving essential health services delivery in future shocks. Separate guides for male and female community members examined access to basic needs including health services during disaster events, coping mechanisms, reliance on informal support systems, barriers to care, and perceptions of state and non-state responses during disasters.

Data collection tools were pre-tested in Thatta by the study team. The pre-test involved staff from a rural health center and a public sector secondary care hospital, along with selected key stakeholders from the PDMA and healthcare providers with prior experience in disaster and emergency management. Feedback from this exercise was used to refine the tools, ensuring greater clarity, contextual relevance, and appropriateness for the diverse respondent groups.

### Data collection process

Data collection was carried out by a team of trained qualitative researchers between August 17^th^, 2020 and October 27^th^, 2020. District-level KIIs and FGDs were conducted in Urdu, while KIIs with provincial and national stakeholders were conducted in English. All interviews and discussions took place in settings designed to protect participants’ confidentiality and were scheduled at times and locations that were convenient for them. Special attention was given to organizing FGDs with community members and LHWs to ensure neutral, accessible venues that supported open and comfortable dialogue.

These sessions were held at Basic Health Units (BHUs) and DHO premises.

To maintain confidentiality and anonymity, each participant was assigned a unique identification code. Written and verbal informed consent was obtained from all KII and FGD participants before data collection began. Verbal consent was collected at the start of each interview or discussion to complement the written consent form, allowing participants to ask clarifying questions and ensuring comprehension, particularly important for individuals with varying literacy levels. When audio-recording was not possible, detailed notes were taken. All transcripts and notes were securely stored and accessible only to authorized members of the research team.

### Data management, triangulation and analysis

Recordings were promptly transcribed and translated into English to preserve accuracy and enable timely analysis. All resulting transcripts and other qualitative data were subsequently imported into MAXQDA for coding and analysis.

A deductive thematic coding approach was employed for data analysis, with the elements of the MAINTAINS conceptual framework for shock-responsive health systems serving as the main codes [33]. Drawing on this conceptual foundation, the coded data was organized into three overarching thematic areas for reporting results:

- Planning, resourcing, and coordination, which captured dynamics within the formal health system and governance structures, including institutional roles, budgeting processes, and coordination mechanisms.
- Service delivery, which examined the system’s capacity to maintain health services provision during shocks.
- Community Resilience and Response, which explored how community health systems and connected sectors contributed to coping strategies, service continuity, and local innovations during floods and droughts.

As for the MIS data analysis, it involved a pre- and post-shock comparison of quarterly DHIS reports from selected districts. For floods, the researchers compared service utilization and morbidity/mortality trends during flood periods with those in the quarters immediately preceding and following floods. For droughts, comparisons were made between prolonged drought-affected quarters and corresponding non-drought quarters.

This approach enabled the team to capture the differential impact of shocks on routine health service delivery. Indicators assessed included immunization services data, outpatient department (OPD) consultations, maternal and obstetric services (antenatal care, deliveries, postnatal care), childhood diseases such as diarrhea and pneumonia in under-fives, selected non-communicable diseases including hypertension and diabetes, and mortality indicators such as maternal deaths, stillbirths, and overall facility-reported deaths.

To enhance the robustness of findings, data were triangulated across respondent types (national, provincial, district, and community levels) and data sources (KIIs, FGDs, and document review).

### Ethical considerations

The study was carried out after obtaining ethical approval from Aga Khan University’s ethics review committee.

## Results

### Planning, resourcing and coordination at national and provincial levels

Disaster planning, resourcing, and coordination in Pakistan’s health system are shaped by institutional structures that differ in their activation and functionality during floods and droughts. Although these structures exist at both national and subnational levels, the below findings suggest that health responses remain fragmented and often reactive with sudden-onset shocks such as floods receiving comparatively more attention than slower-onset shocks such as droughts.

### National and provincial structures

At the national level, three institutions are central to disaster response. The NDMA serves as the overarching multi-sectoral body, supported by its provincial counterparts, the PDMAs. The National Ministry of Health (MoNHSRC) provides technical support to the NDMA during acute shocks such as floods and earthquakes, while the Ministry of Planning and Economic Development plays a coordinating role across all sectors including nutrition, health, WASH, food security, agriculture, and social protection, particularly in drought-affected areas or those with high malnutrition levels [34].

These national structures operate within the framework of Pakistan’s 2011 constitutional devolution, which transferred 21 subjects, including health, to provincial governments. As a result, provincial departments are now primarily responsible for planning, resourcing, and delivering health, nutrition, and water services. The federal government retains a more limited role, focusing on overall coordination, legislation in selected areas, and support to provinces. During acute emergencies such as floods, however, this balance shifts: the NDMA assumes operational leadership at the federal level, while provincial health departments and the MoNHSRC align their activities under the coordination of the NDMA and their respective PDMAs.

Within the MoNHSRC, the National Health Emergency Preparedness and Response Network (NHEPRN) was created in the aftermath of the 2010 floods to consolidate health sector preparedness. However, its role has been limited and largely symbolic, with weak engagement at subnational levels and little influence over provincial planning [20].

### Planning

Weak coordination and capacity of the NEHPRN have constrained the establishment of effective sub-national planning platforms for emergencies and disasters. Further, there have been tensions between the MoNHSRC and provincial departments on planning ownership and role alignment for leading the health response to disasters. Planning mechanisms for floods are comparatively more established than those for droughts. In flood-prone districts – such as Shangla and Dera Ghazi Khan - annual contingency meetings and simulation exercises are conducted under the leadership of PDMA with participation of deputy commissioners, Rescue 1122, and district health teams to plan for resource deployment and evacuation.

*“Plans for floods are already made. Every year we have two mock exercises; in first week of May and first week of June.” (district stakeholder, government)*

However, planning gaps persist. The NDMA’s Multi Hazard Vulnerability and Risk Assessment (MHVRA), used for planning, has been criticized for focusing more on geographical and economic vulnerability while neglecting health service needs and the specific risks faced by vulnerable groups hampering proper planning and response.

*“A vulnerability assessment was conducted by the NDMA. However, a separate report on health was not developed. It was integrated into the broader report, and most of the analysis focused on economics rather than health.” (district stakeholder, international agency)*

Lack of representation of technical health staff within the NDMA and weak coordination with health departments constrain attention to responsive health services and adequate procurement. Preparedness training for health staff on service continuity, reporting, and communication is rare. At the national level, MoNHSRC’s limited capacity and unclear role after devolution constrain its ability to lead. Furthermore, better resourced provinces such as Punjab and Sindh have moved ahead, developing individual crisis response mechanisms for recurrent emergencies, highlighting uneven provincial capacity and fragmented national cohesion. For droughts, anticipatory planning is almost absent.

Droughts are often treated as slow-building phenomena, with planning triggered only once malnutrition or water scarcity reaches crisis levels – hence planning is geared towards mitigating effects of drought rather than preemptive planning for droughts. The Ministry of Planning and Economic Development previously convened multi-sectoral planning through the Scaling Up Nutrition (SUN) Movement, but this weakened after SUN coordination ended [35]. UN agencies are pivotal in disaster response, managing country aid inflows for speedy commodity procurement and distribution, technical assistance, directly managing operations in hard-to-reach areas, and supporting early reporting systems. The UN cluster for humanitarian emergencies supports the NDMA and health counterparts, drawing on the large network of grassroot development NGOs involved with the Polio eradication efforts [36].

### Resourcing

Floods allow for relatively greater flexibility for resource mobilization than droughts. Once emergencies are declared, NDMA and PDMAs can bypass lengthy procurement processes, allowing direct purchases from the market. This enables quicker supply of medicines and relief items, although disbursement delays and supply shortages at district level persist.

*“During emergencies, procurement requirements such as quotations, tenders, and bidding processes are relaxed in accordance with regulations… When an emergency is declared, we [PDMA] are allowed to procure directly from the market, significantly reducing the time usually required for tenders and bidding.” (provincial stakeholder, disaster management)*

Drought-related resourcing is more constrained. Financing relies on formal district requests, often delayed or denied, and incentives such as temporary bonuses for doctors in remote drought-affected areas lack sustainability. Operational costs for outreach, including fuel and transport, were cited as key bottlenecks.

*“Ninety thousand Pakistani rupees were paid to doctors to serve in rural areas during drought, but a sustainable funding stream is lacking.” (district stakeholder, government)*

Across both floods and droughts, disaster-prone districts lack systematic preparedness budgets. Funds are typically triggered during shocks rather than earmarked in advance for training, simulation, or pre-positioning of supplies. Much of the additional international financing is routed through UN agencies, with a considerable share repurposed from development loans rather than new humanitarian aid. Alongside this, philanthropic contributions, corporate social responsibility funds, and religious welfare taxes such as zakat are mobilized during emergencies. However, in the absence of a single consolidated plan and budget, these diverse financial flows result in duplication in some areas, neglect in others, and overall misalignment with actual needs.

Supply chain preparedness further illustrates the contrast between floods and droughts. In flood-prone areas such as Punjab, procurement and pre-positioning of medical flood kits is standard practice, and contingency stocks are sometimes shared between districts through health facility contracting initiatives to external companies such as the People’s Primary Healthcare Initiative (PPHI). Nevertheless, mismatches between actual demand and supply and last-mile delivery challenges remain persistent. In droughts, by contrast, no equivalent forecasting or contingency procurement systems exist, leaving health and nutrition responses largely reactive.

*“Each year in May, medical flood kits are provided to PDMA for further distribution to affected areas.” (district stakeholder, agency)*

*“In this district, contingency items are pre-stocked by PPHI and shared between districts to manage shortages.” (district stakeholder, healthcare provider)*

### Coordination

Coordination mechanisms exist on paper but function weakly across both floods and droughts. During floods, PDMAs and their district counterparts - District Disaster Management Authorities (DDMAs) - are tasked with convening actors across sectors, and deputy commissioners in some districts play an active role. Yet coordination tends to fade after the immediate relief phase. Respondents noted that NDMA and PDMAs approach disaster management primarily through a rescue-and-relief lens, with health often treated as secondary.

*“PDMA is more concerned with rescue and overall measures. We help them in health delivery component during disasters.” (provincial stakeholder, health department)*

In the case of droughts, coordination is even more fragmented than during floods. In provinces where drought has gained political visibility, the Planning and Development Departments have sometimes convened multi-sectoral platforms that bring together health, nutrition, and WASH stakeholders under joint programs. However, such efforts are an exception rather than a rule. In most provinces, coordination has remained largely on paper, with strategies and plans drafted but little evidence of actual implementation or follow-through at the ground level. One of the few entry points for coordinated action has been the issue of stunting, which has drawn attention to drought-affected areas and mobilized multi-sectoral programs. These include nutrition-specific interventions such as monitoring and therapeutic feeding in health facilities, alongside nutrition-sensitive measures such as safe water provision and food supplementation.

The role of non-state actors further illustrates this fragmented coordination across both shocks. While they often mobilize in response to drought, their activities are not systematically integrated into government-led structures, resulting in parallel and poorly aligned efforts. Many operate according to their own mandates. Coordination tends to occur ad hoc through deputy commissioners’ offices during floods, while in droughts engagement is rarer but sometimes formalized through memoranda of agreement [15,37].

*“Currently organizations come with their own mandate but are not filling the gaps rather just completing their own mandates.” (national stakeholder, international agency)*

*“We do not have any specific direct role in any disaster…. We coordinate with local and international NGOs in community mobilization during droughts. We work as a bridge.” (district stakeholder, government social welfare)*

UN agencies play a pivotal role in both floods and droughts, providing technical assistance, managing commodity procurement, and supporting early reporting systems. The UN humanitarian cluster system also leverages grassroots NGO networks, particularly from the polio program [36]. However, respondents emphasized that weak inter-agency coordination and knowledge management means that lessons from each disaster are not systematically embedded into updated contingency plans or standard operating procedures. This reflects a wider national trend: after-action reviews are conducted inconsistently, and when they do take place, their findings and recommendations are rarely disseminated beyond internal circles, limiting their potential to inform future preparedness and response.

*“There are significant lessons to be learnt from each disaster … but the process of learning from successful initiatives have been rather slow.” (national stakeholder, international agency)*

### Service delivery in districts

Service delivery in Pakistan’s disaster-prone areas reveals significant vulnerabilities across health infrastructure, human resources, supplies, and outreach. Yet the nature of these weaknesses diverges sharply between sudden-onset floods and slow-onset droughts as presented below. Analysis of routine health information further confirms these patterns, showing systematic declines in service utilization during both flood and drought periods compared to non-shock years, though with distinct profiles of disruption. Floods, because of their immediacy and visibility, tend to attract emergency responses and international attention [19, 37, 38]. Droughts, in contrast, expose deeper systemic neglect, particularly in remote areas such as Tharparkar, where chronic shortages and weak infrastructure undermine continuity of care. This imbalance has produced stark inequities in access to health services across Pakistan’s provinces.

Floods cause widespread damage to physical health infrastructure. Facilities located along rivers are frequently inundated, forcing the relocation of services into tents, camps, or mobile units. During flood periods, the uptake of acute and emergency curative services at health centers - including outpatient consultations, laboratory investigations, inpatient admissions - was consistently lower than in flood-free months. Primary care centers were more commonly affected due to either direct damage or comprised road access whereas hospitals being more centrally located and equipped with better roads, tended to be more operational.

*“Even the BHUs had been flooded. All the BHUs which were in close proximity to the riverside had been affected too.” (provincial stakeholder, health department)*

Preventive and services such as immunization coverage, pre-and postnatal visits and family planning also dropped. Immunization was offered in camps however fewer pregnant women received the second dose of the tetanus toxoid vaccine and fewer infants completed pentavalent and measles vaccinations. Human resources are mobilized during floods, with additional staff and emergency response teams deployed by both the government and non-governmental organizations. Medical camps are set up quickly, and vaccination services are expanded through government programs, philanthropic groups, and health networks. Mobile units and boats are also deployed to reach isolated populations. Maternal and reproductive health receives particular attention during floods, with efforts to register pregnant women and assign female staff. However, despite these measures, gaps remain in access to services, privacy, and the availability of uninterrupted supplies.

*“Vaccinators don’t get a chance to go there, and they don’t get easy access to it as the river is around 20 kilometers wide… They keep their access through boats.” (district stakeholder, healthcare provider)*

In response, some districts were more successful in establishing contingency measures compared to others, demonstrating uneven response. Mobile medical teams, camp-based services and temporary midwifery stations were deployed, though implementation remains inconsistent and heavily dependent on local capacity. Districts in Sindh province where management of facilities had been outsourced to an independent semi-government company, had more functional 24/7 services as well as upgraded care providing maternal and diagnostic services, even during floods through static centers as well as camps.

*“We have made a cluster of 12 such BHU Plus, where everything related to reproductive health is dealt with. There we have ambulance service; we have a laboratory facility where all the tests related to maternity health are conducted… In floods also we establish camps and continue the antenatal care service. We ensure the immunization rounds as well as family planning in the camps.” (district stakeholder, healthcare provider)*

In contrast, in districts where facilities are managed directly by the government, service continuity is far more constrained. Chronic shortages of anti-snake venom, sterilized delivery kits, and blood products such as fresh frozen plasma limit their ability to respond to acute needs.

*“We get an increase in the cases of insect bites and snake bites. Then in such a situation the fresh frozen plasma in our blood bank become deficit with us.” (district stakeholder, healthcare provider)*

With regards to data, there are multiple reporting systems across various government and civil society providers delivering services during floods, but a central MIS repository has not been developed for flood reporting.

Unlike floods, droughts do not produce immediate destruction but instead expose deep-rooted service delivery weaknesses. Most drought-affected districts are geographically remote, with poorly equipped facilities, long distances to facilities, and inadequate roads and transport. These challenges compound over time. In Tharparkar, drought has been linked to food insecurity, high levels of maternal and child malnutrition, and increased psychological distress.

*“The drought affected millions, resulting in severe food insecurity, poverty, and widespread maternal and child malnutrition… there was also a high rate of suicidal tendencies among women due to the severity of the crisis.” (provincial stakeholder, health department)*

Health needs in droughts are chronic - malnutrition, anemia, and reproductive complications - yet the system offers little continuity or preventive care. Outreach services are rare, and temporary relief is mobilized only when outbreaks occur. Nutrition activities continue to be documented through parallel reporting systems alongside the DHIS, highlighting persistent fragmentation in information flows and coordination. Human resource deployment during droughts remains largely routine, with some surge capacity for nutritional related special activities.

Nutrition services in period of droughts drought typically consist of supplementation programs, including iron, folic acid, and food support for pregnant women. However, coverage is uneven, and without emergency planning, access remains limited and inconsistent.

Despite their differences, floods and droughts share several systemic weaknesses. Essential services such as mental health care, sexual and reproductive health (SRH) are under-prioritized in both contexts and while UN agencies and humanitarian actors have attempted to fill these gaps, interventions are sporadic, rarely integrated into operational frameworks. Although acute illness care is provided to varying extents, sustained care for chronic disease gets more severely disrupted. There is little standardization across providers in terms of which services are to be provided and under which protocols. The qualitative testimonies highlight that while floods produce abrupt service disruptions and short-term collapses in utilization, droughts steadily undermine preventive care and create nutrition challenges. Despite these different pathways, both shocks weaken access to essential services and reveal systemic blind spots, particularly for SRH.

*“Disasters have a profound adverse impact on the SRH of women, yet SRH remains inadequately addressed in most situations in Pakistan. While humanitarian agencies and UN bodies have responded to SRH needs during droughts and floods, a more systematic and sustained approach is urgently needed.” (provincial stakeholder, civil society organization)*

### Community resilience and response

Communities in Pakistan’s flood and drought affected regions have demonstrated resilience under pressure, improvising to meet urgent needs amidst chronic state neglect. This broad pattern, which is further detailed in the findings that follow, is not without limits. It is shaped and constrained by long-standing structural vulnerabilities, such as landlessness, indebtedness, gender inequality, inadequate public services, and fragile infrastructure, that precede disasters and determine how different households experience and respond to shocks. The nature of the shock also plays a critical role: while floods bring abrupt and visible destruction, droughts gradually erode livelihoods and well-being over time. In both contexts, communities lean heavily on informal support systems, drawing on kinship ties, collective action, and local innovation. Yet, these coping strategies are unevenly distributed and often insufficient in the face of recurring and large-scale crises.

### Livelihood losses and resource scarcity

Floods and droughts have severely disrupted local economies, leading to the loss of crops, livestock, and other essential livelihood assets. In flood-prone districts, damage to land and standing crops left many families unable to repay loans taken to cultivate leased land.

*“The rains destroyed all the cotton in 2020. It was perfectly ready and then the rain came. Now we are in debt and have been asking the debtor for some time. Some people are selling their animals to cover these debts.” (male community member)*

The loss of livestock - an important source of both income and nutrition - further undermined household resilience. With milk production disrupted, even basic dietary needs became difficult to meet.

*“We produce milk here from our buffaloes and animals, but we sell it to big dairy farmers like Nestle. So now we have to buy this powdered milk because we cannot consume our own milk. We cannot meet our dietary needs.” (female community member)*

In drought-affected areas, crop failure and barren grazing lands forced families to migrate or sell animals they had accumulated over years, undermining long-term food and economic security.

*“The grass ends, and all our animals are hungry… we migrate to a different city which is at a higher altitude.” (male community member)*

*“All the crops are gone, animals gone… We remain hungry for days.” (male community member)*

Loss of livelihoods further exacerbated food insecurity and restricted access to basic services, including healthcare. Food scarcity led to significant reductions in meal frequency and nutritional intake, especially among women and children. Basic resources such as cooking fuel and clean water were also compromised. In rural districts, where most households rely on stored wood for cooking, floods rendered this supply unusable, forcing families to burn unsafe materials. To navigate these challenges, communities frequently turned to kinship networks and local solidarity mechanisms. Family members, especially those in cities, provided food or lent resources through informal systems such as *lain dain* (mutual loans). Neighbours contributed by sharing essentials like milk and water, while shopkeepers often extended credit for basic goods like flour. These hyperlocal safety nets were essential for daily survival.

*“No outside organization really helps us, we mostly rely on each other within the family. When someone is in need, we sell our animals to support one another.” (male community member)*

*“Like I said, only small acts are possible in a place like this, we help each other with things like sugar or atta [flour]. The shopkeeper sometimes gives us goods on credit, and our relatives step in with small loans when they can.” (female community member)*

### Water access and community-led initiatives

Access to clean water was another serious vulnerability, worsened by both floods and droughts. Inundated wells and contaminated runoff in flood-hit areas led to widespread illness.

*“Because of the water lots of illnesses happen … Our kids, women and elderly are all dying… We are not getting any aid… I often feel like I will hit someone.” (female community member)*

Despite limited means, communities came together to address these water challenges. In drought-affected areas, they raised funds or contributed livestock to deepen wells.

Though the water retrieved was often brackish and unsafe, it remained the only option.

Some villages constructed protective walls around wells, while others built local water connections. Community-based organizations also played a critical role in mobilizing resources for WASH projects.

*“We collected a small contribution from everyone in the village. For those who couldn’t pay in cash, we asked for a baby goat instead, which we could sell to cover their share. That’s how we managed to dig six to eight water points. Then we approached the Thardeep Rural Development Program, and they responded by installing solar pumps instead of hand pumps.” (male community member)*

*“After the flood, Al Falah organization went door to door collecting money from community members to restore our local water supply scheme. Beyond that, they routinely stepped in to repair any damage whenever needed.” (male community member)*

### Community repair of infrastructure and housing

In the absence of state support, communities also repaired critical infrastructure. They built flood barriers, restored washed-out roads, and constructed temporary bridges to ensure access to nearby areas.

*“I took the initiative to build a flood barrier. We rented a bulldozer and used the front blade of a tractor to construct it. Everyone contributed some money to help. It held back water during several floods, but it finally gave way during the 2010 flood.” (male community member)*

*“During the 2020 flood, a road in Damorha was washed away. But people immediately came together and repaired it enough to make it passable. They even built makeshift bridges to restore access.” (male community member)*

Flooding also devastated housing infrastructure, displacing families and forcing them to seek shelter in precarious and unsafe locations. Makeshift dwellings on roadsides exposed residents - especially women, children, and persons with disabilities - to heightened protection risks.

Despite such hardship, communities continued to extend solidarity, even in times of grief. During deaths or personal tragedies, neighbours collectively supported funerals and helped navigate logistical barriers caused by floods.

*“When a poor man in our village passed away, the whole community came together to support his family. We helped evacuate the floodwater, cleared the paths, and arranged the coffin and burial.” (male community member)*

### Access to healthcare

Access to healthcare emerged as one of the most pressing and persistent challenges during both floods and droughts. These shocks exposed serious deficits in the accessibility, affordability, and quality of public health services. In many cases, health facilities were either entirely absent or severely dysfunctional. Where facilities did exist, they were often understaffed and provided substandard care, leading to widespread distrust and avoidance among community members. Health outreach services, such as mobile medical camps and nutrition programs, were similarly criticized for their ineffectiveness and lack of real support. Many viewed these efforts as symbolic or performative, failing to address actual needs.

*“The Sehat people reach camps but there are no medicines, it’s just a show piece. Regardless of what the disease is, we will get paracetamol.” (male community member)*

Affordability remained a major barrier to accessing care. With livelihoods loss, crop income diminished, and livestock sold, families struggled to cover even basic transportation costs or user fees.

*“I had to give Rs 3000 to a nurse so that she could deliver a child. The hospital would not take her and told her to go away to Multan. What can a person do in this situation? The nurse took Rs 3000 and delivered the child on the streets. Poor people do not have Rs300, how will they pay Rs 3000?” (male community member)*

Amid this absence of reliable state services, communities organized their own health access mechanisms. Local health workers and volunteers stepped in to provide essential support such as carrying the sick or pregnant to higher ground or arranging rotating access to limited mobile medical teams across neighbouring villages. These grassroots efforts were vital, particularly in areas that were physically cut off due to flooding.

*“I even had a patient who had been very serious, helped her cross the way by having her lifted on shoulders and that was how she got her medicine.” LHW*

Healthcare accessibility challenges led to delays in seeking care, with many residents turning to informal providers or spiritual healers or undertaking arduous journeys often on motorcycles or foot to access help. In some cases, this resulted in dangerous outcomes, such as home births in high-risk situations.

### Gendered burdens and psychological toll

Despite some evidence of resilience, it remained uneven. Households with fewer assets, women-headed families, and persons with disabilities were more likely to be excluded from communal support systems. Gendered impacts were particularly severe. In both floods and droughts, women bore the brunt of unpaid labour, caregiving, and logistical burdens. In drought-affected areas, the loss of donkeys - sold to repay debts - meant women carried heavy clay pots for kilometres each day to fetch water. The psychological toll was heavy for both women and men. Stress, food insecurity, displacement, and overwork contributed to reported cases of depression, aggression, and suicidal ideation. Women shared stories of fainting from hunger or fatigue, and some communities reported suicide by poisoning particularly among young women.

*“The women have also lost their minds because of the stress. We have many cases of suicide. They drink some sort of poison” (male community member)*

In summary, community resilience in Pakistan’s flood and drought affected regions is obvious, rooted in mutual aid, social networks, and collective effort. Yet these informal systems are fragile and insufficient substitutes for systemic support.

#### S3. Results summary

### Discussion and conclusions

This study provides a rare, multi-level perspective on Pakistan’s response to climate-related shocks, contrasting floods and droughts across national, provincial, district, and community levels. By analyzing both the formal health system and community coping strategies, it demonstrates that resilience is shaped not only by institutional planning, resources, and coordination, but also by the ways households and local networks adapt under pressure.

At the institutional level, Pakistan has developed a comprehensive disaster management architecture comprising of the NDMA and PDMAs supported by the UN cluster and several planning documentations but despite more than a decade of successive shocks and UN agencies support, preparedness and response into the health sector remains fragmented and largely reactive.

Floods due to their acute nature and visibility receive greater attention and resources than droughts, however longer-term preparedness and resilience planning for both shocks is weak. Evidence suggests these structures prioritize visible rescue and relief efforts over sustained service continuity, echoing earlier critiques from Pakistan and global literature [11,17]. Comparable studies in sub-Saharan Africa similarly show that risk assessments emphasize infrastructure and economic losses while neglecting health service vulnerabilities and equity considerations [12]. Coordination over planning, resourcing and operationalization emerges as a critical weakness between health agencies and disasters agencies, as well as between government health agencies, private health providers and civil society organizations. These patterns reflect broader challenges across low- and middle-income countries, where weak governance, poor intersectoral coordination, and chronic underinvestment limit the adaptive capacity of health systems [2,20].

Differences between floods and droughts are particularly marked in planning and financing. Floods trigger contingency meetings, simulation exercises, and rapid mobilization of humanitarian financing, though coordination often remains weak [21].

Droughts, by contrast, are slow-onset, attract little political salience, and rarely prompt anticipatory planning or flexible financing - a pattern mirrored in other low- and middle-income countries where droughts are systematically underprioritized [19]. This imbalance highlights how the visibility of a shock, rather than its long-term impact, drives the allocation of resources and political attention, reflecting on deep political economy dynamics. As elsewhere, shocks that are acute, visible, and disruptive tend to attract donor and government attention, while slow-onset crises such as droughts remain neglected. This visibility bias undermines equity, as the poorest and most marginalized — especially in drought-affected areas — face cumulative losses without systematic support.

Service delivery challenges illustrate the dual weaknesses of hardware (infrastructure, workforce, supplies) and software (coordination, standardization, reporting). In floods, acute curative care, vaccination, and temporary maternity care services are deployed through mobile units, medical camps, vaccination campaigns on boats but unevenly, whereas ongoing care needs such as for chronic illness, mental health and family planning are less well provided. Human resources are relatively quickly mobilized but constrained by the shortage of products, protocols, standardized delivery packages and streamlined monitoring metrics and systems. In drought-affected districts, structural weakness of remote health systems is exacerbated with under-nutrition and WASH issues requiring upskilled staff, commodities, integrated reporting systems and sustained community-based outreach.

Ineffective response deepens household poverty, as families sell assets, incur debt, and face potentially catastrophic out-of-pocket costs - an effect widely observed across low and middle-income countries [2]. The consequences for maternal, newborn, and child health are particularly severe. Pakistan already ranks among the poorest performers globally for maternal mortality and child malnutrition [39,40], with stunting levels above the Asian average [41,42]. Climate shocks aggravate these burdens by disrupting newborn and maternity care, limiting contraceptive access, and heightening food insecurity. A rapid assessment conducted in Pakistan found that 84% of women in flood camps were dissatisfied with relief services, 77% reported no access to SRH kits, and 69% of adolescent girls stopped schooling after floods [43]. These statistics underscore how SRH and maternal and child health remain systematically neglected in disaster response, despite their centrality to population resilience.

At the community level, resilience is evident but fragile. Overall, systematic planning and coordination systems to involve the communities in preparedness and response monitoring are missing, delegating communities to passive recipients larger than building resilience. Moreover, despite the presence of an extensive national social protection database in Pakistan there has been little attempt to link risk assessment, preparedness and response with social protection databases. Households rely on kinship networks, migration, asset sales, and collective infrastructure repair - strategies consistent with adaptive behaviors in other fragile settings [15,44]. However, these coping mechanisms are unevenly distributed, inequitable, and unsustainable. Women bear disproportionate burdens of caregiving, water collection, and psychological distress, while persons with disabilities and poorer households are often excluded from community support. This highlights the need for gender-sensitive and equity-focused approaches that recognize community capacities without over-relying on them as substitutes for systemic support.

Taken together, these findings underscore the persistent gap between Pakistan’s disaster management architecture and the lived realities of affected communities. Bridging this gap requires moving from reactive, event-driven interventions to integrated resilience strategies. International frameworks, such as the WHO’s operational framework for climate-resilient health systems, call for embedding preparedness into routine planning, ensuring flexible financing for anticipatory action, and institutionalizing after-action reviews to foster learning [1,36]. At the operational level, district health offices should lead capacity-building for frontline staff, focusing on continuity of antenatal, postnatal, growth monitoring, maternity, and family planning services during shocks. Working in close coordination with civil society and development partners, district authorities must also develop actionable strategies to prioritize vulnerable populations and ensure essential health care reaches those most at risk.

While this study focused primarily on preparedness and response, future resilience also depends on recovery and transformation - leveraging crises to strengthen governance, expand community-based models such as the BHU Plus, and integrate health into broader climate adaptation strategies. Persistent gaps remain, including the neglect of continuity of services such as for mental health, chronic disease, family planning, and the weak use of routine health information during shocks and fragmented reporting. Addressing these gaps is critical not only for Pakistan but also for other climate-vulnerable low- and middle- income countries, where building climate-resilient health systems requires sustained, equity-sensitive, and evidence-driven approaches. Future practice and research should explore how resilience can be institutionalized across recovery phases, with greater emphasis on health-specific operational planning, stronger coordination and role alignment, clear specification of standardized service packages and protocols, robust monitoring metrics, and proactive planning and monitoring for community-level responses.

### Strengths and limitations

This study has several strengths. First, it provides a rare, multi-level perspective on health system resilience to climate shocks in Pakistan, integrating national, provincial, district, and community-level insights. By triangulating data from KIIs, FGDs, and document reviews, the analysis offers a rich and nuanced understanding of both formal system responses and community coping mechanisms. Second, the study was conducted by a multi-disciplinary team of researchers, which helped reduce positionality bias in the analysis and interpretation of findings. The use of the MAINTAINS conceptual framework also provided a systematic lens for examining health system resilience across multiple building blocks and phases of shock response.

At the same time, some limitations should be acknowledged. On the participant side, as most respondents were selected through purposive and snowball sampling, there is a risk of over-representation of individuals already engaged in formal disaster response structures, potentially limiting the diversity of perspectives. Community participants were drawn from high-risk districts accessible within security clearances, which may have excluded some of the most vulnerable or marginalized populations, particularly in conflict-affected areas of Balochistan and western Sindh.

## Data Availability

The data underlying this article cannot be shared publicly to safeguard participant privacy. De-identified data may be obtained from the corresponding author upon reasonable and justified request.

## Acknowledgments

Acknowledgement to Zarak Hussein Ahmed, Sana Hyat, for their support in fieldwork and to Shama Dossa, Minhaj Kidwai, Zafar Fatmi for insightful inputs to the Pakistan work.

## Supporting information captions

**S1. Documents guiding disaster response and areas for consolidation**

**S2. MAINTAINS program’s framework**

**S3. Results summary**

---

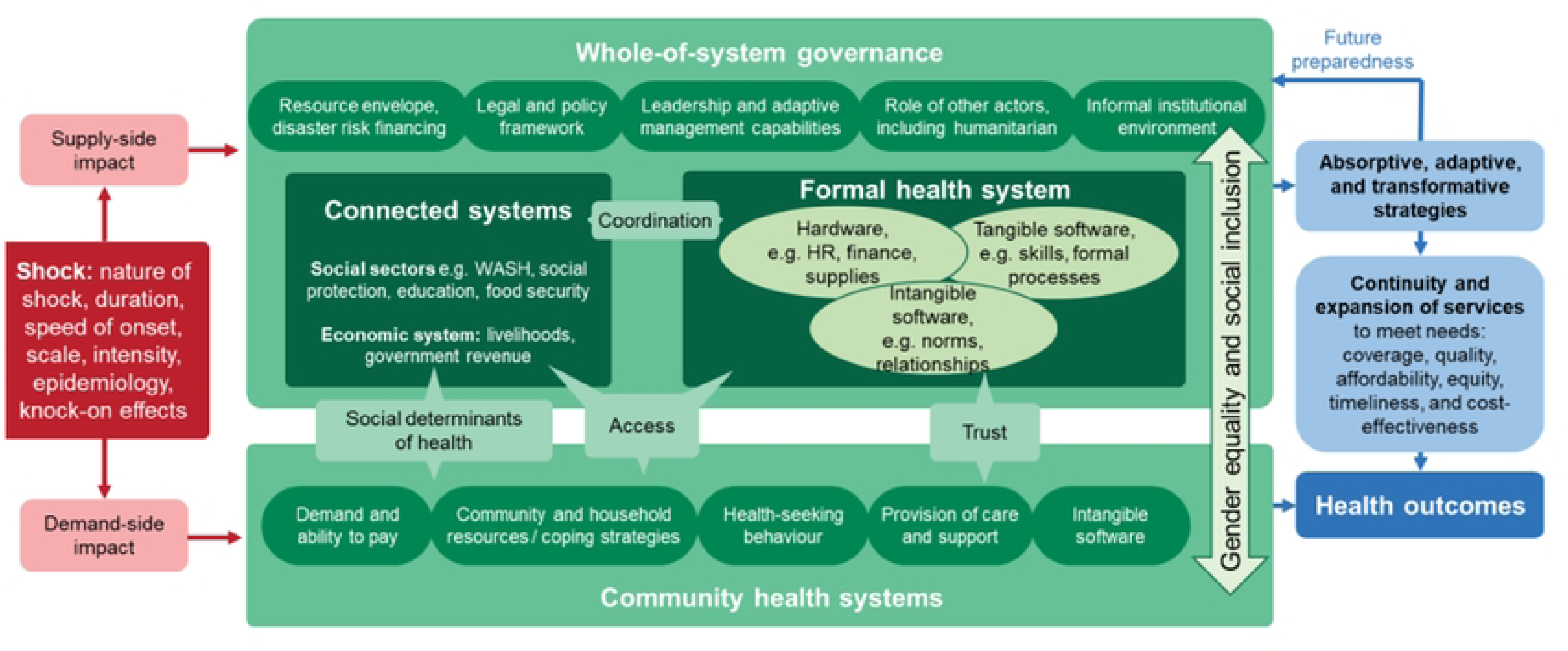

